# The landscape of structural variants in male infertility identified by optical genome mapping

**DOI:** 10.64898/2026.02.27.26347236

**Authors:** Anja Kovanda, Alenka Hodžić, Urška Kotnik, Tanja Višnjar, Rebeka Podgrajšek, Aleksander Andjelić, Helena Jaklič, Aleš Maver, Luca Lovrečić, Borut Peterlin

**Affiliations:** Clinical institute of genomic medicine, University medical Centre Ljubljana, Ljubljana, Slovenia; Faculty of Medicine, University of Ljubljana, Ljubljana, Slovenia; Department of Human Reproduction, Division of Obstetrics and Gynecology, University Medical Centre Ljubljana, Ljubljana, Slovenia

**Author notes:** corresponding author: prof. Borut Peterlin.

**Keywords:** optical genome mapping, structural variants, copy number variants, male infertility, amplification, complex variants, exome sequencing

## Abstract

**STUDY QUESTION:** [Do structural genomic variants, that can be identified by using optical genome mapping, contribute to male infertility?]

**SUMMARY ANSWER:** [By using optical genome mapping we can identify several types of structural variants, both known and new, that may contribute to male infertility.]

**WHAT IS KNOWN ALREADY:** [Traditional approaches such as karyotyping, *CFTR* and chromosome Y microdeletion testing are successful in explaining clinical findings in ∼30% of MI patients, leaving the rest without a genetic diagnosis. Recent research suggests at least 265 genes may play a role in male fertility. While the assessment of the roles of copy number variants and single nucleotide variants in monogenic forms of disease in these genes is underway, much less is known about structural variants.]

**STUDY DESIGN, SIZE, DURATION:** [We performed a longitudinal case/control study on a total of 220 individuals; 88 patients with male infertility, negative for cytogenetic abnormalities using karyotyping, and molecular testing for chrY microdeletions, and *CFTR* gene variants, and 132 healthy male individuals that underwent optical genomic mapping for other reasons. Exclusion criteria for the control cohort were low-sperm quality and/or inclusion in IVF procedures. The study was approved by the National Medical Ethics Committee of the Republic of Slovenia (reference number: 0120-213/2022/6). Optical genome mapping was performed from an aliquot of whole blood collected for routine testing purposes at the Clinical Institute of Genomic Medicine (CIGM), UMC Ljubljana from January 2023 to November 2024.]

**PARTICIPANTS/MATERIALS, SETTING, METHODS:** [We examined structural variants in 220 participants by using optical genome mapping, which was performed with DLE-1 SP-G2 chemistry and the Saphyr instrument. The de novo assembly and Variant Annotation Pipeline were executed on Bionano Solve3.7_20221013_25 while reporting and direct visualization of structural variants was done on Bionano Access 1.7.2. All obtained variants were filtered using the Bionano Access software and in-house generated gene/regions of interest panel bed files. The first filter was applied to include variants below a population frequency of 10%, and overlapping the regions of interest. Subsequently, all variants occurring with frequency 0% in the internal manufacturer variant dataset were manually evaluated for possible involvement of the overlapping genes or regions in biological processes involved in MI. The male infertility cohort also underwent research whole exome analyses as previously reported. All results of optical genomic mapping were confirmed by an appropriate alternative method where available.]

**MAIN RESULTS AND THE ROLE OF CHANCE:** [We show that the overall number of structural variants in MI patients does not differ from that of healthy individuals. By looking in detail at genes and regions associated with MI, we identified 21 rare variants absent from controls in 25.0 % of MI patients, of which five were likely causative, and two would be missed by using traditional approaches. These variants include inversions, duplications, amplifications, deletions (e.g. *SPAG1*), and insertions/expansions (e.g. *DMPK*), that were validated using additional methods. While the remaining SV cannot be currently classified as pathogenic according to existing criteria, they open a new avenue in genetic research of MI.

**LARGE SCALE DATA:** [Variants reported in this study were deposited into ClinVar under accession numbers SUB15650956 (https://www.ncbi.nlm.nih.gov/clinvar/)]

**LIMITATIONS, REASONS FOR CAUTION:** [Technical limitations of optical genome mapping include the lack of DLE-1 labelling of centromeric and telomeric regions, the inability to detect Robertsonian translocations, the unclear exact location of smaller structural variants located between the DLE-1 labels, and unclear boundaries in case of their location in segmentally duplicated regions (this limitation is shared with other methods). The ACGM criteria of rarity are also hard to apply, as the fertility status of the individuals in healthy population databases such as GnomAD and DGV is unknown. Similarly, gene-associated phenotype and the proposed inheritance model both need to be considered as parts of the ACMG criteria, but for many candidate genes associated with MI, no model of inheritance has yet been proposed.]

**WIDER IMPLICATIONS OF THE FINDINGS:** [Currently, with the established diagnostic approaches we are able to resolve ∼30% of male infertility cases, with ∼70% of patients remaining undiagnosed. The significance of our work is in showing that rare structural variants can be identified in MI, by using optical genome mapping, opening new avenues of research of the genetics of this important contributor to human fertility.]

**STUDY FUNDING/COMPETING INTEREST(S):** [All authors declare having no conflict of interest in regard to this research. This work was funded by the Slovenian Research and Innovation Agency (ARIS) Programme grant P3-0326: Gynecology and Reproduction: Genomics for personalized medicine]

**Lay summary:** Male infertility affects about 5% of adult males and has complex causes, including genetic ones, such as mutations in the *CFTR* gene, small deletions on chromosome Y, and balanced translocations, but currently we can only find a genetic cause in ∼30% of patients. This means ∼70% of cases remain undiagnosed but potentially, they too may have a yet unknown genetic cause. Indeed, so far research has shown at least 265 genes have been proposed to play a role in male fertility. In these genes, there has so far been limited research of single nucleotide variants and of copy number variants, but many structural variants are not visible using commonly used methods in clinical genetic testing. Therefore, apart from chromosome Y microdeletions and chromosomal numerical and structural anomalies, such as balanced translocations, the role of smaller structural variants in male infertility is unknown, but based from what we know from other diseases, they also may play a role in male infertility.

Optical genome mapping is a novel method for the detection of structural variants, such as balanced and unbalanced translocations, insertions, duplications, deletions, and complex structural rearrangements in a wide range of sizes. By using optical genome mapping to test a cohort of 88 infertile men and 132 healthy controls, we aimed to provide the first insights into the range of SV that may be associated with MI. We found, by using optical genome mapping, the overall number of structural variants in MI patients not to be significantly different to the control group. However, by looking at genes and regions associated with MI, we can find rare structural variants that are absent from controls in 25.0% of MI patients. These variants include inversions, duplications, amplifications, deletions (e.g. deletion in *SPAG1*), and insertions/expansions (e.g. in *DMPK*), that were validated using additional methods. Five of these variants (5.6%) were likely causative, and two would be missed by traditional approaches. While the remaining SV cannot be currently classified as pathogenic according to existing criteria, they open a new avenue in genetic research of MI.

## 5. Introduction

The prevalence of male infertility (MI) is estimated to be between 2.5 and 12% in different populations and has a complex aetiology including environmental and genetic contribution (Leslie *et al*., 2024). Genetic testing is indicated in cases with non-obstructive azoospermia, oligozoospermia and nonpalpable vas deferens, and according to the guidelines includes karyotyping, testing for chromosome Y (chrY) microdeletions (for primary infertility, azoospermia, and oligozoospermia), and cystic fibrosis transmembrane conductance regulator (*CFTR*) gene testing (for vasal agenesis or idiopathic obstructive azoospermia) (Brannigan *et al*., 2024; Minhas *et al*., 2025). While the yield of the traditional testing approaches ∼30%, the causes of infertility in the remaining patients remain unknown. So far over 250 genes have been suggested to play a role in male fertility, with only a few found to have clinical significance so far (Podgrajsek *et al*., 2025a). Currently, the role of single nucleotide variants (SNV) and copy number variants (CNV) in these candidate genes is under active investigation (Oud *et al*., 2025; Podgrajsek *et al*., 2025b). However, there is yet no information on many other types of potentially causal structural variants (SV) that are not visible using next generation sequencing or microarrays. While chrY deletions and translocations have a known role in MI, the role of smaller, complex and balanced SV that are below the 5-10 Mb resolution of karyotyping, remains to be determined.

Optical genome mapping (OGM) is a novel method based on fluorescent labelling of DNA motifs, and the imaging of individual, labelled, high-molecular weight DNA molecules, with subsequent mapping to reference DNA. This enables the detection of SV, such as translocations, insertions, deletions, and complex structural rearrangements in a wide range of sizes, likely to be missed by the technologies currently in routine use (Dremsek *et al*., 2021; Mantere *et al*., 2021).

Our aim was to comprehensively evaluate the landscape of structural genomic variants by using OGM, in order to examine their possible etiological contribution to MI.

## 6. Materials and methods

### Participants

A total of 95 patients were referred for genetic testing due to MI at the Clinical Institute of Genomic Medicine (CIGM), UMC Ljubljana from January 2023 to November 2025. OGM patients represented a subset of the previously described exome sequencing (ES) male infertility research cohort (Podgrajsek *et al*., 2025b) (Supplement 1).

The comparison cohort consisted of 132 healthy male individuals of whom 45 were healthy fathers (HF) and 87 were healthy males whose partners had recurrent miscarriages (RM), and underwent OGM testing for other reasons. Exclusion criteria for the comparison cohort were low sperm-quality and/or inclusion in IVF procedures.

### Sample inclusion and prior genetic testing

As a standard approach all 95 patients with MI included in the study first underwent genetic testing using karyotyping, chrY microdeletion testing (Devyser AZFv2 kit, Stockholm, Sweden), and *CFTR* gene variants (Pelzman and Hwang, 2021). Patients in whom a genetic cause was previously identified using the standard approach were excluded from the study, while in new patients the standard approach was performed in parallel with the OGM analyses. In this way OGM was performed on 88 MI patients in total.

### Ethics declaration

The study was approved by the National Medical Ethics Committee of the Republic of Slovenia (reference number: 0120-213/2022/6)(Podgrajsek *et al*., 2025b). Written informed consent for OGM and WES testing, granting the Clinical Institute of Genomic Medicine (CIGM), rights to publish the findings of genetic testing in de-identified form in scientific literature, was obtained from all individuals included in the study at their clinical visit, and all procedures were performed in line with the Helsinki declaration and all relevant National guidelines.

### Optical genome mapping (OGM)

OGM was performed as previously described (Rogac *et al*., 2023). Briefly, high-weight molecular DNA was extracted from 1.5 million lymphocytes from whole blood (EDTA collected) using the SP Blood & Cell Culture DNA Isolation Kit or the SP-G2 Blood & Cell Culture DNA Isolation Kit following manufacturer instructions (Bionano Genomics Inc., San Diego USA). The following day, DNA molecules were labeled with the DLE-1 enzyme using the Direct Label and Stain (DLS) Kit or Direct Label and Stain-G2 (GLS-G2) kit (Bionano Genomics Inc.). Labeled DNA was loaded on the three-flowcell Saphyr Chip® G2.2 or G2.3 (Bionano Genomics Inc.) and ran on the Saphyr instrument (Bionano Genomics Inc.) to reach a minimum yield of 500 Gbp (DLE-1 label, [GRCh38] reference genome). The de novo assembly and Variant Annotation Pipeline were executed on Bionano Solve3.7_20221013_25 while reporting and direct visualization of structural variants was done on Bionano Access 1.7.2.

### Genes and regions of interest

Genomic locations on the [GRCh38] human genome to be examined as regions of interest were generated using the UCSC Table browser tool from a large gene panel containing research genes associated with MI. 265 genes were included in the generation of regions of interest: *ADAD2, ADCY10, ADGRG2, AK7, AKAP9, ALG13, AKAP4*, *AMELY, AMH, AMHR2, AR, ARL2, ARL2BP, ARMC2, ART3, ASZ1, ATG4D, ATM, AXDND1, BCORL1, BEND2, BRCA2, BRD2, BRDT, BRWD3, C11ORF80, C14orf39, CAMK4, CATIP, CATSPER1, CATSPER2, CCDC115, CCDC146, CCDC155, CCDC34, CCDC39, CCDC40, CCER1, CD1D, CD63, CDC20, CDK5RAP2, CEP128, CEP131, CEP70, CEP78, CFAP206, CFAP43, CFAP44, CFAP47, CFAP54, CFAP61, CFAP69, CFAP70, CHD5, CHD7, CIMIP4, CLCA4, CREM, CSTF2T, CT55, CTCFL, DAZL, DDX25, DDX3Y, DDX53, DHRSX, DMC1, DMRT1, DNAH1, DNAH17, DNAH2, DNAH6, DNAI1, DNALI1, DND1, DNHD1, DNMT3A, DNMT3B, DRC1, DZIP1, ELMO1, ESR2, ESX1, EXO1, FAM47B, FAM47C, FAM50B, FAM9B, FAM9C, FANCA, FANCB, FANCM, FBXO43, FKBP6, FKBPL, FMR1NB, FOXP3, FSHB, FSIP2, GALNTL5, GCNA, GMCL1, GPR143, GTF2H3, HAUS7, HENMT1, HFM1, HIPK4, HORMAD1, HSD17B4, HSF2*, *HYAL3, IFT140, IGSF1, IHO1, INHA, INSL3, KASH5, KATNAL2, KCND1, KCTD19, KDM3A, KIAA1210, KISS1R, KLK14, LHB, LHCGR, M1AP, MAGEB4, MAGEB6, MAGEE2, MAJIN, MAP3K15, MAP7, MBOAT1, MCM8, MCM9, MCMDC2, MEI1, MEIOB, MLH1, MLH3, MMRN1, MND1, MNS1, MOV10L1, MSH4, MSH5, NALP14, NANOS1, NANOS2, NEURL4, NPAS2, NR0B1, NR5A1, NUP210L, ODF4, PDHA2, PGK2, PHF7, PICK1, PIWIL1, PIWIL2, PKD1, PLCZ1, PLK4, PMFBP1, PNLDC1, POC1B, POLR2C, PRM1, PSMC3IP, QRICH2, RAD21L1, RAD51AP2, RBBP7, RBMXL2, RBMXL3, REC114, REC8, REDIC1, RHOXF1, RHOXF2, RHOXF2B, RNF212, RNF212B, ROS1, RPL10L, RXFP2, SECISBP2, SEMA5A, SETX, SGO2, SHOC1, SLC22A16, SLC26A8, SOHLH1, SPAG17, SPATA22, SPATA3, SPATA6, SPIDR, SPINK2, SPO11, SPOCD1, SSX3, STAG3, STRA8, STX2, SUN1, SYCE1, SYCE1L, SYCP1, SYCP2, SYCP3, TACR3, TAF4, TAF4B, TAF7L, TBC1D25, TBCCD1, TDRD6, TDRD7, TDRD9, TDRKH, TEKT3, TENT5D, TERB1, TERB2, TEX10, TEX11, TEX12, TEX13A, TEX14, TEX15, TKTL1, TMPRSS9, TOP6BL, TOPAZ1, TRIM37, TRIM71, TTC21A, TTLL9, UPF2, USP26, USP9Y, UTP14C, VCX3A, VHL, WDR66, WNK3, WT1, XRCC1, XRCC2, ZCWPW1, ZFAND3, ZFPM2, ZFX, ZFX, ZMYM3, ZMYND15, ZNF318, ZNF85, ZPBP*, and *ZSWIM7*.

Additionally, the [GRCh38] genomic locations of the AZFa, AZFb and AZFc regions of interest and the locations of Deyser probes for Y microdeletion assay regions were added to the MI regions of interest bed file (Supplementary information). The following regions/probes mapping to the chrY were added: AZFa (Deyser basic probes sY86 sY625 sY84 M259), AZFb (P5/proximal P1, sY134), AZFb (Deyser basic probes sY127 sY131 sY134), AZFc type b2/b4 (sY254 sY255), AZFc (Deyser basic sY254 sY255 sY157), Gr/Gr (SY1291 absent sY1191 present, includes DAZ, BPY2, CDY1 1, 6 Mb), Yp11.3 (Deyser basic probes sY14 ZFX/Y), Yq11.21 (Deyser basic probe sY81), Yq11.221 (Deyser basic probe sY90), sY100 (DYS196 SHGC-5482), sY105 (DYS201 SHGC-5487), sY1064, sY1065, sY116 (DYS208 SHGC-5492), sY1182, sY1191 (DYS11), sY1191 (G73809), sY1192 (G67166), sY121 (DYS212 SHGC-5496), sY1224 (G72342), sY127 (DYS218), sY1291 (G72340), sY131 (DYS222 SHGC-5505), sY134 (DYS224 SHGC-5506), sY14, sY143 (DYS231), sY153 (DYS237), sY157 (DYS240 SHGC-5512), sY158 (SHGC-5513), sY160 (tandem DYZ1 AY598347), sY254 (tandem), sY255 (Deyser basic probe for DAZ1-4), sY255 (tandem), sY625 (G65849), sY81 (SHGC-5469), sY82 (SHGC-5470), sY84 (DYS273), sY86 (DYS148), sY88 (DYS276), sY90 (DYS278 SHGC-5477).

Finally, the variants below 0% prevalence in the internal Bionano database were checked manually for overlap with genes or regions that may be associated with MI.

### Identification of SV of interest

All obtained variants were filtered using the Bionano Access software and in-house generated gene/regions of interest panel bed files. The first filter was applied to include variants below a population frequency of 10%, and overlapping the 265 gene panel and the AZFa, AZFb and AZFc regions of interest. Subsequently, all variants occurring with frequency 0% in the internal OGM dataset (Bionano Genomics, Bionano Solve Theory of Operation; CG-30190) were manually evaluated for possible involvement of the overlapping genes or regions in biological processes involved in MI.

### Variant interpretation

We report only those genomic variants that have statistical support based on the adequate genomic coverage and chosen analysis type for the detection of: copy number change, deletion, insertion, duplication, inversion, intra- and inter-chromosomal translocations, as determined by internal Bionano QC parameters. The method does not enable the analysis of regions that do not contain DLE-1 labelling sites (centromeres, telomeres, and other heterochromatin regions). The assessment of variants as SV of interest was performed according to the ACMG in ClinGen guidelines (Riggs *et al*., 2020), by taking into account the following databases: GnomAD – (https://gnomad.broadinstitute.org/), Database of genomic variants (DGV) – (http://dgv.tcag.ca/gb2/gbrowse/dgv2_hg19/) (MacDonald *et al*., 2014) DECIPHER (https://www.deciphergenomics.org/), ClinVar (https://www.ncbi.nlm.nih.gov/clinvar/), and ClinGen (https://dosage.clinicalgenome.org/). For evaluation of SV we used adjusted criteria based on the most likely effect of the variant based on CNV and SNV classification criteria (Richards *et al*., 2015; Riggs *et al*., 2020). Because of less available information on SV (compared to CNVs), the result can be less informative (please see Limitations).

### Limitations

Our work has several inherent limitations, which are shared with other similar studies involving patient and comparison group composition, and limitations of variant classification. Several inherent aspects of MI genetics further complicate a straightforward classification of SV in case of this diagnosis. The relatively high prevalence of this condition in the general population (estimates differ for various counties and range between 2.5 and 12.0% (Leslie *et al*., 2024)) and the lack of information on fertility of the healthy controls included in the population databases such as GnomAD and DGV means we cannot easily apply the criteria of rarity when assessing variants. Similarly, because of anatomical and biological differences, while gene-associated phenotype and the proposed inheritance model both need to be considered as parts of the American College of Medical Genetics (ACMG) criteria, for many candidate genes associated with MI, there yet may not be a known model of inheritance. Currently it is therefore not possible to finally resolve classification of variants in case of heterozygous variants in genes with proposed autosomal recessive inheritance with MI with additional clinical features, as the heterozygous variants, while may confer only an increased risk or present an incompletely penetrant phenotype.

Additionally, the remaining technical limitations of OGM concern the lack of DLE-1 labelling of peri-centromeric and telomeric regions. This affects the method’s ability to detect Robertsonian translocations and other translocations between such regions, that do affect the success of reproduction. Similarly, OGM faces similar challenges to sequence-based methods in forming correct assemblies across regions spanning segmental duplications, e.g. on chrY, where the ambiguity of mapping can result in several alternative assemblies.

Interpretation of identified SV remains difficult both because of limited knowledge of known normal OGM genetic variation at the moment, and the difficulty in applying ACMG and the joint consensus of (ACMG) and the Clinical Genome Resource (ClinGen) (The ACMG Laboratory Quality Assurance Committee *et al*., 2015; Riggs *et al*., 2020) recommendations to the many different SV such as balanced translocations, inversion and complex SV detected by OGM, where the sequence or the exact breakpoints may remain unknown. Many specific SV lack clear guidelines for classification, and so the interpretation of these SV need to be carefully considered on a case-by-case basis. This means many identified variants at the moment remain variants of unknown significance, and in order to also exclude possible technical artefacts in some cases, whenever possible, the results should be confirmed by using an independent method. While for many rare-disease cases a trio setup is preferable to resolve causality, in case of MI this may not be feasible. Hopefully, some of these challenges may be overcome in time through the establishment of a larger database of OGM detectable normal human genomic variation.

### Variant validation

Variants were validated by additional methods depending on variant type. Deletions on chrY were validated using the Devyser AZF Extension Multiplex Polymerase Chain Reaction kit. Large inversion breakpoints were confirmed using long-range PCR amplification across the putative breakpoints, and intragenic CNVs were confirmed using exome sequencing as previously described (Podgrajsek *et al*., 2025b).

### Figure preparation

All figures were prepared from original visualizations generated by Bionano Access 1.7.2 software (Bionano). The Integrative Genomics Viewer (IGV) was used to visualize the raw WES data (Robinson *et al*., 2011). The UCSC Genome Browser Viewer was used to visualize the region of interest in the context of neighboring genomic regions (Nassar *et al*., 2023). The composite figures were technically prepared in terms of size, layout, format and type of file with no modification to original data, from the original visualizations, by using GIMP 2.10 (The GIMP Development Team, 2019).

### Data availability

Variants reported in this study were deposited into ClinVar under the accession number SUB15650956 (https://www.ncbi.nlm.nih.gov/clinvar/). The datasets generated during and/or analyzed during the current study are not publicly available due to the possibility of containing additional potentially sensitive genomic information. The datasets are available from the corresponding author on reasonable request.

## 7. Results

We performed OGM on a total of 220 participants: 88 MI patients and 132 control males.

On average, MI patients had 4220 SVs (clustered counts from MQR) of which 1270 (30.09%) were deletions, 2832 (67.11%) insertions, 61 (1.44%) duplications and 53 (1.26%) inversions. This distribution did not differ significantly from the control group, where on average 4322 SV per patient were observed: 1318 (30.49%) deletions, 2887 (66.79%) insertions, 58 (1.34%) duplications, 55 (1.27%) inversions (unpaired t-test).

Among these variants, we evaluated rare variants overlapping the 265 genes and other known regions involved in MI (please see Methods section for further details), which we defined as variants of interest.

In this way we identified a total of 31 rare SV in 44 of the 88 patients (50.0%), and 9 such rare SV in 23 of the 132 controls (17.4%) which was statistically significant (Χ^2^= 26.455, p< 0.0001).

In total, 22 rare SV were identified only in the MI group, in a total of 22 of the 88 patients (25.0%). Two patients had more than one variant of interest, and one variant was found in two independent patients (Table 1,2, and 3).

**Table 1:** Hemizygous structural variants in male infertility patients.

**Table 2:** Large structural variants in male infertility patients.

**Table 3:** Small intragenic structural variants in male infertility patients.

Of these 22 rare SV of interest, 10 were located on the sex chromosomes (Table 1), and 12 on the autosomes, of which 4 SV were large and above 100 kb (Table 2), and 7 were small and below 100 kb in size (Table 3). While the large SV contained several genes, the small SV were mostly intragenic (Tables 2 and 3).

Among the MI unique variants, five variants in five patients were shown as likely causative (5.6%; 5/88), using additional testing approaches (Tables 1, 2, and 3). The first was ogm[GRCh38] 8q22.2(100,138,992_100,172,318)×1, a 12.1 kb deletion in a 33.3 kb region, that includes exons 1-2 of the *FBXO43* gene (OMIM:609110, NM_001029860.4), the *POLR2K* gene (OMIM:606033), and the first three exons of the *SPAG1* gene (OMIM:603395, NM_003114.5) (Figure 1). Biallelic pathogenic variants in the *FBXO43* gene represent a known cause of spermatogenesis defect type 64 (OMIM:619696), while biallelic pathogenic variants in the *SPAG1* gene represent a known cause of primary ciliary dyskinesia type 28 (OMIM:615505), manifesting with situs inversus and infertility (Sironen *et al*., 2020). Similar pathogenic deletions (ClinVarID: 88681) in this region have previously been described in a compound heterozygous state with pathogenic SNV in the *SPAG1* gene, in the context of primary ciliary dyskinesia, that includes MI (Knowles *et al*., 2013; Djakow *et al*., 2016; Boaretto *et al*., 2016). Indeed, by additional WES testing, we confirmed the presence of this deletion to be in the *SPAG1* gene. WES analysis also identified a second pathogenic SNV variant, which was consistent with infertility, and the secondary finding of situs inversus, in our patient.

**Figure 1:**
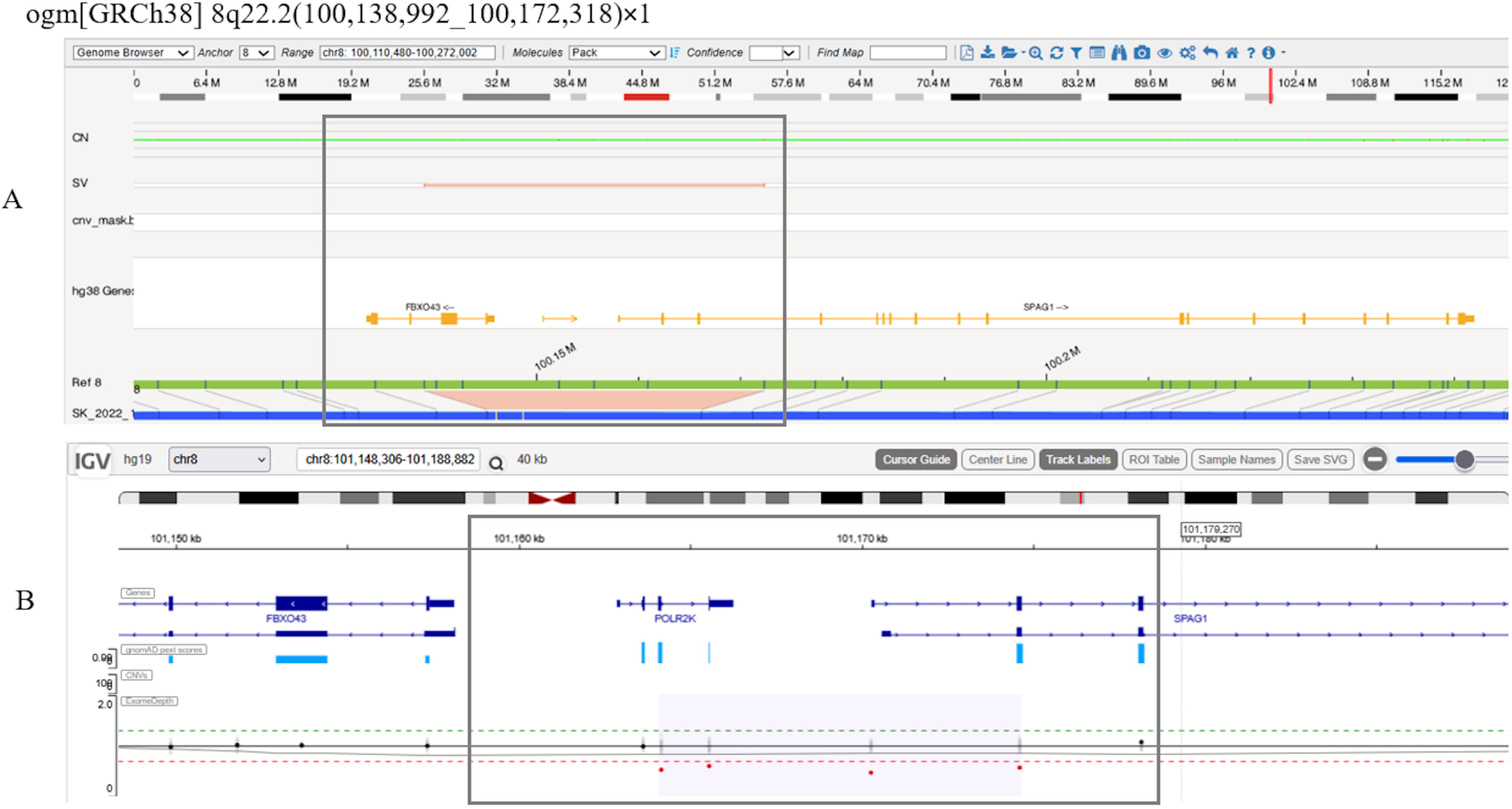
Deletion in *SPAG1,* identified using OGM and confirmed using WES. A: Bionano Access software viewer displaying OGM result in the target region (GRCh38). B: IGV Browser viewer of WES sequencing result displaying coverage and sequences in the target region (hg19).

The second causative SV was initially identified using OGM as a 1.3 kb insertion within the 19.4 kb region, ogm[GRCh38] der(19;?)ins(19)(q13.32;?)(45,752,584∼45,771,947;?), containing the 3’ UTR region and exons 11-15 of the *DMPK* gene (OMIM:605377, NM_004409.5), the SIX5 gene (OMIM:600963), and exons 6-15 of the *MEIOSIN* gene (NM_001310124.2) (Figure 2). Heterozygous expansions of CTG triplets in the 3’UTR region of the *DMPK* gene are associated with autosomal dominant hereditary myotonic dystrophy type 1 (OMIM:160900, GeneReviews: NBK1165). Depending on the size of the expansion, the clinical presentation of myotonic dystrophy type 1 can vary from mild clinical myotonia, which may include MI (Pan *et al*., 2002; Kim *et al*., 2012; Puy *et al*., 2020), to congenital myotonic dystrophy. As OGM cannot detect the nucleotide sequence, we performed WES and targeted *DMPK* expansion analyses. The OGM identified insertion was visible on WES as an expansion located in the intronic region of the *DMPK* gene (Figure 2). The additional targeted testing confirmed this SV as a pathogenic *DMPK* expansion, which was consistent with the main presentation of infertility and clinically mild myotonic dystrophy in the patient.

**Figure 2:**
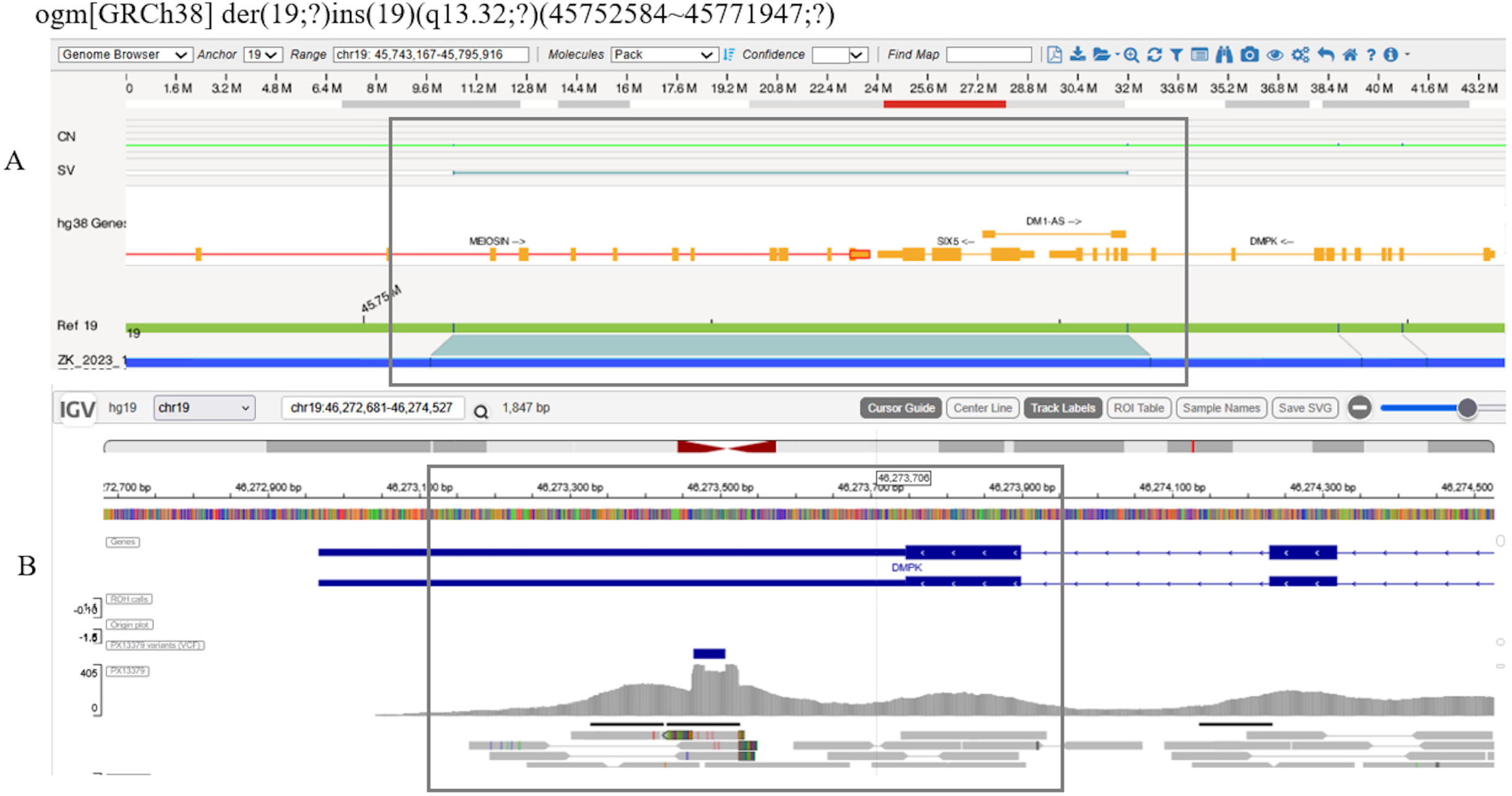
Pathogenic expansion in *DMPK*, identified as an SV using OGM and confirmed using WES and target molecular testing. A: Bionano Access software viewer displaying OGM result in the target region (GRCh38). B: IGV Browser viewer of WES sequencing result displaying coverage and sequences in the target region (hg19).

Finally, we also found three chrY microdeletions that were initially missed by standard chrY microdeletion testing (Supplemental information).

A further 16 SV of interest, potentially associated with MI were found in 18/88 patients (20.4%) of patients (Table 1, 2, and 3). These SV consisted of inversions (355.7 kb in Xq28, and 8.6 Mb in 20q11.23q13.12), insertional translocations (13q31.1), and complex duplications (429.8 kb in 2p25.1, 4.451 Mb in 7p12.1p11.2, and 364.1 kb in 7q35) or amplifications (123.5 kb, 505.2 kb in Xq24, Xq26.3) that were reported for the first time and are unlikely to be detected using classical MI testing approaches. The Xq28 inversion occurred in 2 patients.

The smaller SV we found were also unlikely to be detected using the currently recommended methods in use for MI (Table 3), but most could be confirmed using WES. Among the variants identified among both patients and cohorts (SI Table 1, Supplemental Information), the chrY microdeletions and duplications in the regions Yq11.223, Yq11.23, Yq11.223q11.23 (Navarro-Costa *et al*., 2010; Yu *et al*., 2015; Colaco and Modi, 2018), as well as the ∼579.5 kb amplification in 10q11.22, were the most common SV observed (SI Table 1). In two patients and two controls we identified the AZFc gr/gr deletion risk factors.

These variants have been observed previously, but none were significantly enriched in MI patients compared to healthy controls. 3 rare SV were identified in homozygous form only in the MI group, with heterozygotes identified in both the MI group and the controls (Table S1). The presence of most of the remaining SV of interest was also confirmed using additional technology (See Methods, Supplemental information), and their possible association with MI is further explored in the discussion.

## 8. Discussion

By performing OGM and analyzing rare structural variants in 265 genes and other known regions involved in a total of 220 individuals, we were able to detect 5 likely causative SV in 8.0% (5/88) of MI patients as well as further 21 rare SV of interest that were detected only in the MI cohort.

Two of the likely causative SV, deletion in *SPAG1* (Figure 1) and an expansion in *DMPK* (Figure 2), were unlikely to be detected using standard testing approaches in evaluation of MI, but required additional molecular testing in order to confirm the diagnosis.

The interpretation of the clinical implications of the remaining SV potentially associated with MI requires further functional characterization, as the existing guidelines for variant interpretation define criteria for SNV and CNV that may not be applicable in case of all SV (The ACMG Laboratory Quality Assurance Committee *et al*., 2015; Riggs *et al*., 2020) (See Methods, Limitations section).

Unsurprisingly, 34.6% (9/26) of all rare unique SV were found on the sex chromosomes, including the chrY microdeletions in conformations initially missed by standard molecular microdeletion Y testing (Supplemental information). However, because of the high-repetitiveness sequence of the chrY and the OGM resolution in these regions, it is currently not yet possible to determine the exact breakpoints with confidence in all cases by any of the available technologies. Therefore smaller SVs potentially covering *RBMY1A1* and *RBMY1B* (Yan *et al*., 2017), *SPRY3* (OMIM:300531), *VAMP7* (OMIM:300053), and *ILR9* (OMIM:300007) (Tannour-Louet *et al*., 2014; Chávez-López *et al*., 2020; Ponomarenko *et al*., 2020) (Li and Hamer, 1995; Helena Mangs and Morris, 2007) remain candidates or controversial candidates only. Of note is that, the telomere-to-telomere (T2T) assembly of chrY has only very recently been made, using a combination of several available technologies (Rhie *et al*., 2023), and has highlighted many challenges of this repetitive chromosome. Hopefully the further improvements of the OGM and WES pipelines of assembly, based on this groundbreaking work, will translate to a better resolution of similar SV in the future. Similarly, the duplications and inversions identified on chromosome X, currently remain variants of unknown significance (further discussed in the Supplementary information).

The three types of identified SV on the autosomes, whose role remains to be elucidated were a.) duplications and amplifications with unknown location in the several hundred kb to several Mb range, b.) large inversions in the several Mb range (Table 2), c.) small insertions and d.) small deletions below the size of 100 kb. The detailed interpretation and appearance of variants from Tables 1–3 is given in the Supplementary information.

The large duplications, amplifications and an inversion we identified are unlikely to be found by karyotyping, due to its detection size limitations. Of note is that SV in this size range are usually located between smaller segmental duplication sites, therefore their boundaries or edges may remain unresolved regardless of the technology used.

Most of the remaining SV identified by OGM were small deletions and insertions (Table 3). Because of the methodological limitations of OGM, the exact location of these variants within larger DLE-1 labelled intervals are unknown, requiring additional WES or targeted molecular testing. Interestingly, some of these deletions were not called by WES analysis pipeline possibly due to intronic location of the breakpoints, but after their identification using OGM, we were able to confirm their presence upon manual inspection of raw WES data (Supplementary information). On the other hand, we also found two homozygous intronic variants, that were could not confirm using WES due to lack of intronic coverage (Supplement) in candidate MI genes. Apart from one small SV found in both the MI and comparison cohort, and two homozygous variants from the MI cohort found in heterozygous form in the comparison cohort, the remaining 7 small intragenic variants were absent from the comparison cohort.

We found heterozygous deletions, confirmed by WES, in the *CLCA4, NEDD4, DNAH9, CFAP52*, and *HAUS1* gene in which biallelic pathogenic variants have been associated with MI by several independent studies (Frühmesser *et al*., 2013; Ta-Shma *et al*., 2015; Zhou *et al*., 2017; Dougherty *et al*., 2020; Hodžić *et al*., 2021; Tang *et al*., 2021; Zou *et al*., 2022; Cheung *et al*., 2023) (Supplemental information). For all of them comparable deletions could also be found in the reference databases in rare individuals with unknown fertility status. However, as we were unable to find additional pathogenic SNVs in these genes through WES analysis, these heterozygous variants currently cannot explain the MI phenotype.

In regard to insertions, apart from the likely causative expansion in *DMPK* described in the results, we found variants of interest in *MRE11* and *TSPB1*, genes that have also been implicated in MI by several studies (Petrini *et al*., 1995; Nolan *et al*., 2004; Su *et al*., 2007; Zhao *et al*., 2012; Bonache *et al*., 2014; Liu *et al*., 2017; Hu *et al*., 2020), but we were unable to confirm their exact location or composition using WES (Supplement).

Our results show that OGM, in addition to detecting known SV previously associated with MI, such as chrY microdeletion, can also detect rare small SV affecting candidate MI genes that may be missed by WES, as well as medium size complex variants of unknown significance, that are currently invisible using other technologies.

Despite better technical resolution in identifying SV, an important limitation of our study is the difficulty in the clinical interpretation of SV, because a.) the existing variant interpretation guidelines may not be applicable in case of all SV (The ACMG Laboratory Quality Assurance Committee *et al*., 2015; Riggs *et al*., 2020), b.) the reference variant databases containing variants from the general population do not yet contain many SV, with OGM SV being particularly rare, and c.) the relatively high incidence of MI, together with the fertility status being mostly unknown for the healthy individuals included in the reference databases, means the criteria of the variants’ rarity are harder to apply. Finally, despite the use of stringent exclusion criteria, the possibility remains that some of our comparison group males may unknowingly be affected by subfertility, since the condition is so common. Nevertheless, we show that by using OGM we can identify additional SV contributing to the complex aetiology of MI.

## Conclusions

By using OGM we show that patients with MI do not have an inherently higher number of SV than healthy controls and that structural rearrangements are a rare occurrence. By looking more specifically at rare SV in genes and regions associated with MI, we were able to identify additional two rare likely causes of MI that would be missed by traditional approaches. The remaining SV that were absent from the control group are also likely to be missed using traditional approaches and consisted of duplications and amplifications with an unknown location, inversions and small intergenic deletions and insertions. While at the moment, we are unable to assign causality to the such rare SV, our finding open further research into genetics of MI.

## 9. Author’s contribution

AK and BP conceptualized and designed the study. AK, UK, TV, RP, AA, HJ, AM, LL, and BP contributed to the acquisition of data, performed the analysis, and performed interpretation of data. AK and BP drafted the article and all authors revised it critically for important intellectual content. All authors approved the final version to be published. All authors are accountable for all aspects of the work.

## 10. Acknowledgements

We profoundly thank the patients and controls participating in this study. We also thank Ms. I. Anžel Lara and Ms. S. Žitko for their excellent technical assistance.

## 19. Supplementary Data

Supplement table 1: Structural variants in male infertility patients and controls.

Supplementary data.pdf

Supplement bedfile.txt

