## Supplemental file for "The landscape of structural variants in male infertility identified by optical genome mapping"

**Supplemental information**

The identified SV of interest from Tables 1,2,3 and SI Table 1 are given in their genomic order.

**ogm[GRCh38] 1p22.3(86,565,210_86,582,137)×1**

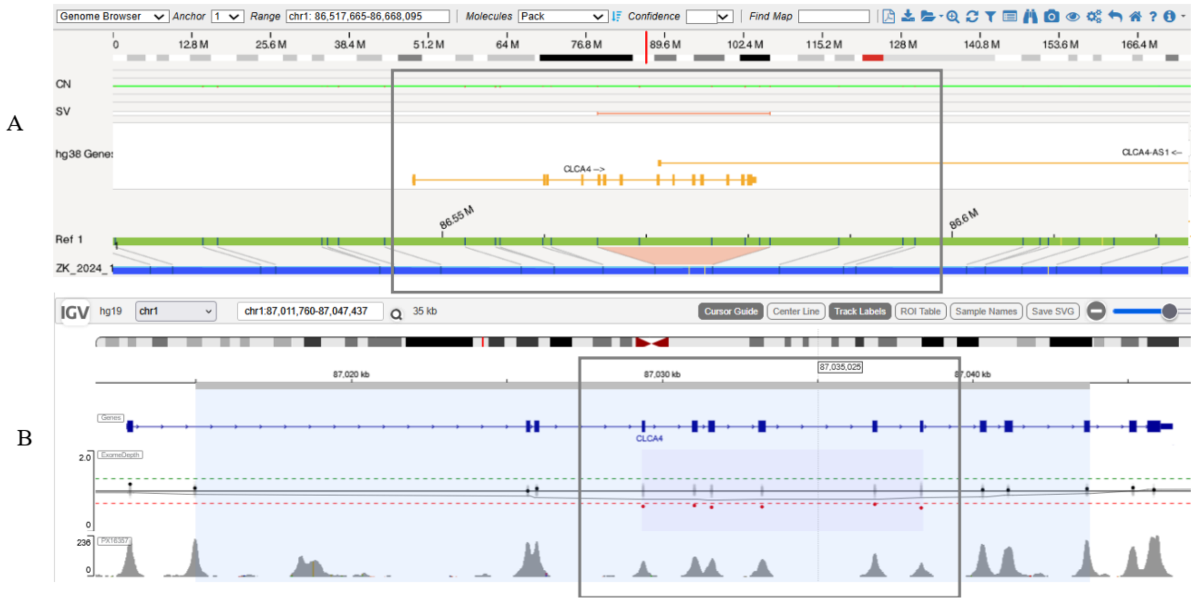


This SV is a 11.3 kb heterozygous deletion located in the 16.9 kb region containing exons 4-14 of the *CLCA4* gene. Biallelic pathogenic variants in the *CLCA4* gene (OMIM:616857) have been reported to be associated with idiopathic MI by two independent studies, including ours (*1*, *2*). We found an overlapping heterozygous deletion of similar size, defined as a loss of function variant, to be present in the GnomAD database (DEL_CHR1_4299F19D) in males with a frequency of 0.003.

**ogm[GRCh38] 2p25.1(10,440,150_10,869,905)×3**

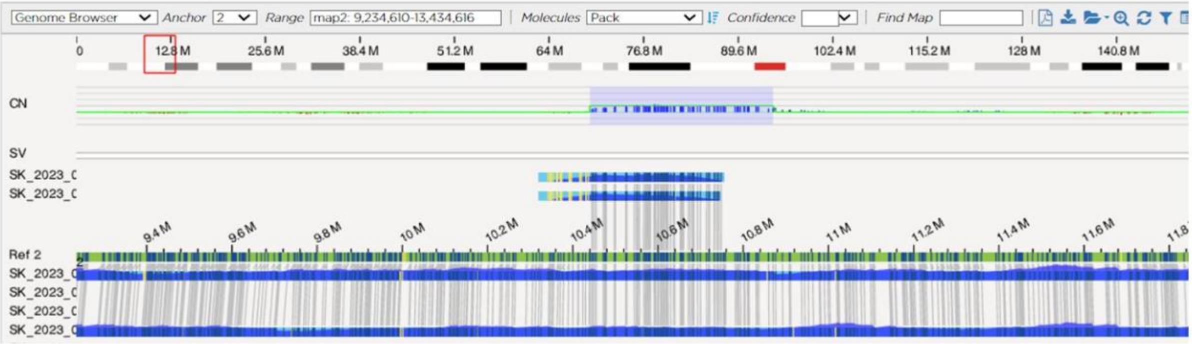


The 429.8 kb gain, ogm[GRCh38] 2p25.1(10,440,150_10,869,905)×3, contains the genes *ATP6V1C2* (OMIM:618070), *NOL10* (OMIM:616197), *ODC1* (OMIM:165640), *PDIA6* (OMIM: 611099), of which none were previously associated with MI, and similarly to other complex duplications in this size range, could not be confidently located due the absence of any DLE-1 sites from the duplication boundary region.

**ogm[GRCh38] der(6)ins(6;?)(p21.32;?)(32,354,608~32,357,248;?)**

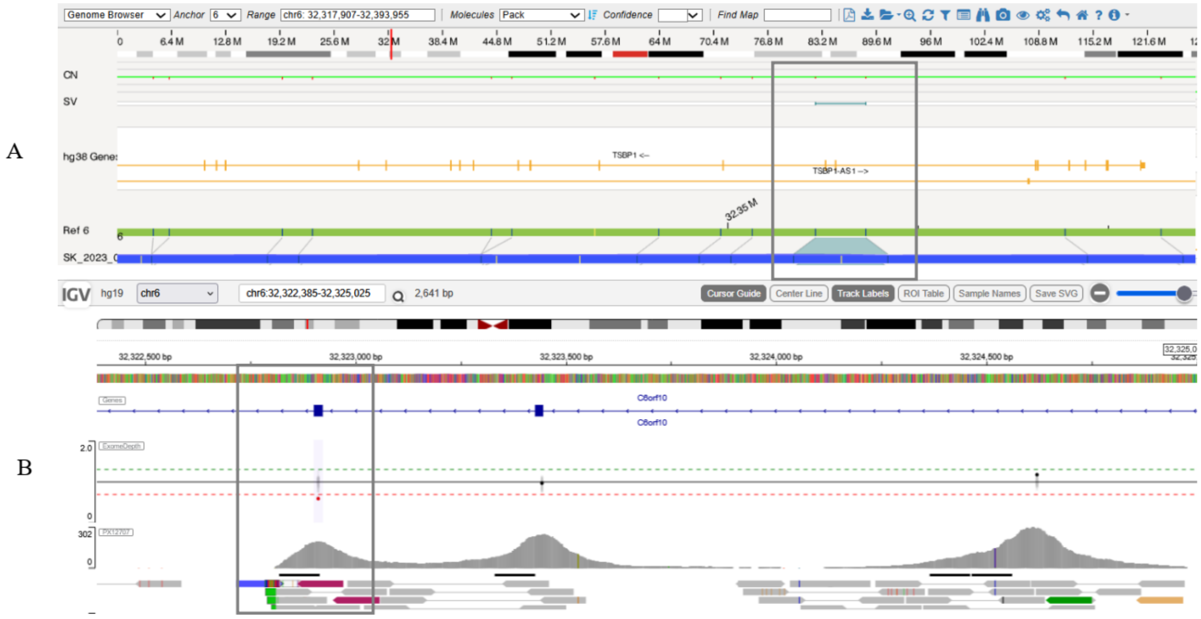


The homozygous insertion, ogm[GRCh38] der(6)ins(6;?)(p21.32;?)(32,354,608~32,357,248;?), was 2.3 kb in size in a 3.6 kb region, which includes the exons 7 and 8 of the testis-specific binding protein 1 (*TSBP1)* gene (NM_001286474.2). TSBP1 (OMIM:618151) has the highest expression in the testes, where it is supposed to stimulate protein kinase A activity, which in turn is crucial for successful sperm motility (*3*, *4*), with previous studies proposing its involvement in MI (*5*, *6*). No similar insertions were found in the reference databases, however, in our study, an insertion of the same size and at the same location was present in heterozygous form in six other MI patients, as well as 9 comparison cohort individuals. WES showed the homozygous insertion is intronic, however its sequence is not identical to the heterozygous insertion in the other individuals (Supplement, Figure X), despite the appearance of being the same size using OGM analysis, which will be explored further in future studies.

**ogm[GRCh38] 7p12.1p11.2(53,576,787_58,027,799)×3**

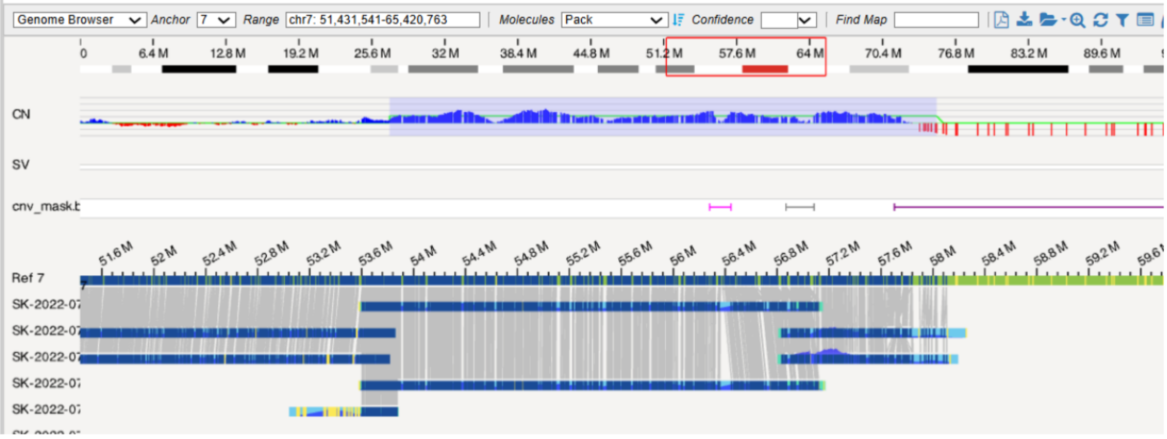

The location of the 4.5 Mb gain, ogm[GRCh38] 7p12.1p11.2(53,576,787_58,027,799)×3 (Table 2, Supplement), could not be resolved with certainty, leaving open the possibility of a cryptic insertional translocation or a marker chromosome, since this SV is located right next to the centromere. Various reciprocal translocations of chromosome 7 have previously been associated with MI (*7*, *8*). Unfortunately, we were unable to confirm the location of this variant using FISH, because of loss of patient to follow-up.

**ogm[GRCh38] 7q35(143,516,254_143,880,379)×3**

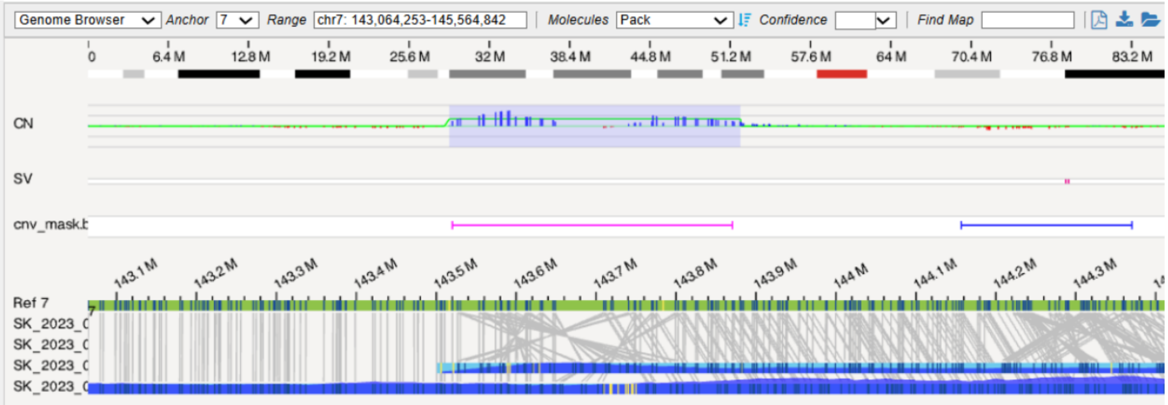


The 364.1 kb gain ogm[GRCh38] 7q35(143,516,254_143,880,379)×3 does not have clear boundaries enabling confident localization of the duplication, leaving open the possibility of its involvement in insertional translocation at an unknown (OGM and karyotype invisible) location. While such SV may not necessarily affect the health of the carrier, their implications for fertility remain to be determined.

**ogm[GRCh38] 10q11.22(46,100,908_46,680,405)×4~5**

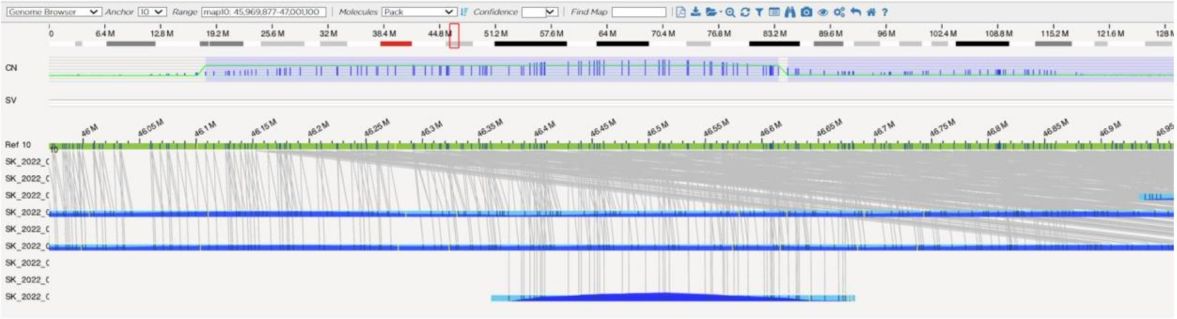


The amplification ogm[GRCh38] 10q11.22(46,100,908_46,680,405)×4~5 is notably located in a region where the hg19 and GRCh38 significantly differ, and mapping between the two assemblies is not clear-cut.

While at least two independent studies have previously found the 10q11.22 region as duplicated in studies of MI (Halder et al., 2017; Kikas et al., 2023), the duplications of this region, not amplifications, may be present in healthy subjects (with unknown fertility status) quite commonly (DGV Gold, gssvG4289, gain of 0.15% frequency, European and African population). A 100kb smaller duplication has also been identified in homozygous form in Europeans quite frequently (GnomAD SV v4.1.0, DUP_CHR10_52F15DB9, 0.27 allele frequency, European population). Yet, to the best of our knowledge, we are the first to report an amplification in this larger region, and interestingly, the 10q11.22 region contains the genes *PPRY1* (OMIM:601790), *GPRIN2* (OMIM:611240), *SYT15* (OMIM:608081), and the *ANTXRL* gene (HGNC:27277), of which the latter is exclusively expressed in the testis (Fagerberg et al., 2014). Hopefully, the possible significance of this region may be resolved further with improved T2T assembly and annotation of chromosome 10 in the in the future.

**ogm[GRCh38] der(11;?)ins(11)(q21;?)(94,455,649~94,461,741;?)**

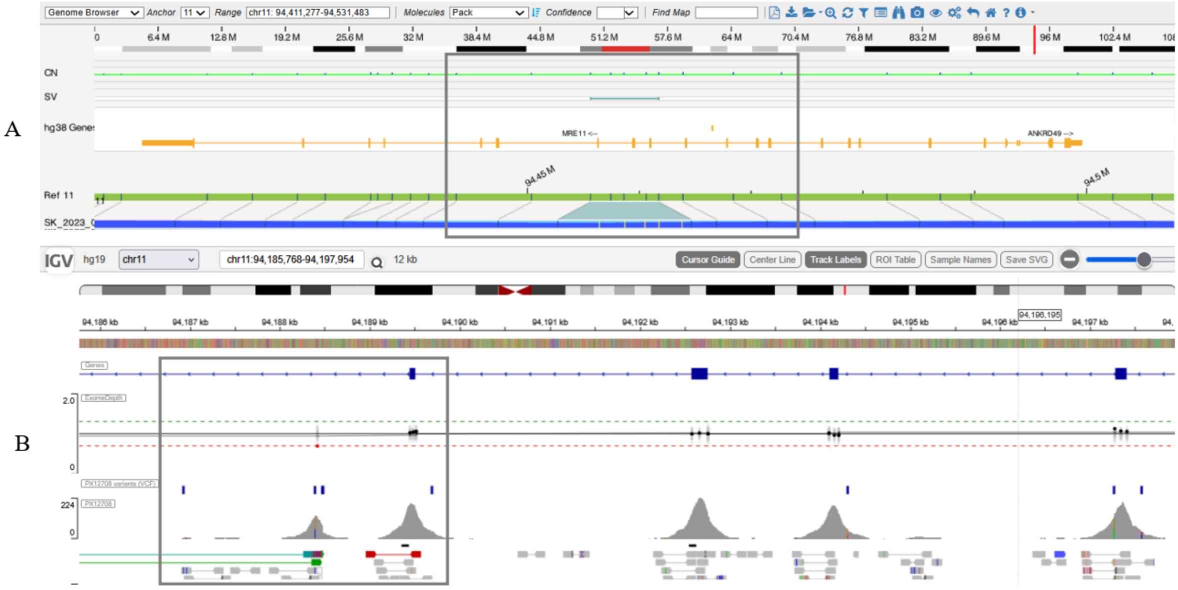


The intragenic insertion, ogm[GRCh38] der(11;?)ins(11)(q21;?)(94,455,649~94,461,741;?), was 5.8 kb in size in a 6.1 kb region of 11q21, containing exons 12-14 of the *MRE11* gene (NM_005591.4). Pathogenic homozygous or compound heterozygous variants in the *MRE11* gene are associated with autosomal recessive ataxia-telangiectasia-like-disease 1 (OMIM:604391). Functionally, the gene is involved in the repair of double-stranded DNA and is expressed mainly in rapidly dividing tissues, its expression is high in the testis (*9*). Studies studying mRNA and protein expression show MRE11 is reduced in patients with non-obstructive azoospermia (*10, 11*). The variant did not appear in any of the reference databases. We were unable to accurately determine the location of this variant in the WES data suggesting it likely represents an intronic variant.

**ogm[GRCh38] 12q23.1(99,598,042_99,615,914)×0**

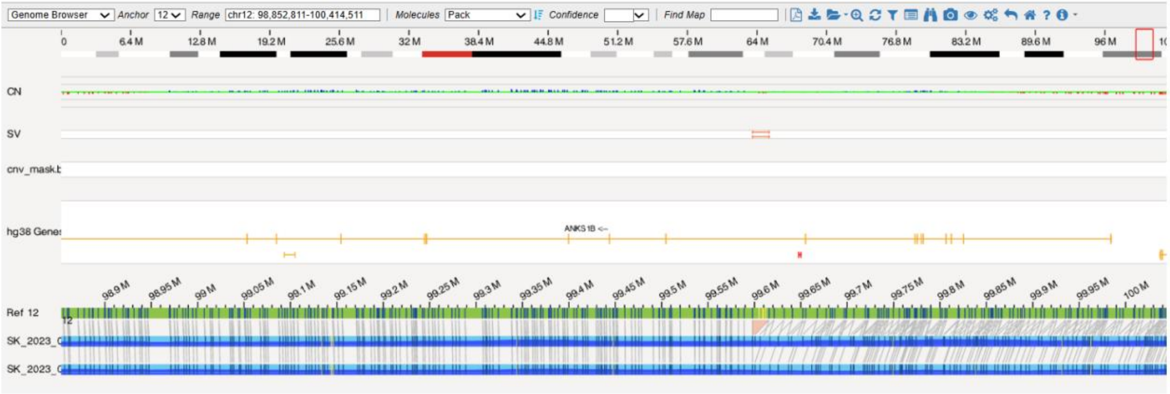

The homozygous deletion, ogm[GRCh38] 12q23.1(99,598,042_99,615,914)×0, was 15.2 kb in size and located within a 17.9 kb region of intron 9 of the *ANKS1B* gene (OMIM:607815, NM_001352186.2). In previous reports, a heterozygous deletion within intron 9 of the *ANKS1B* gene, 27.6 kb in size was found associated with MI (*12*, *13*), however heterozygous deletions of similar size in intron 9 are common in the general population, and so the contribution of this variant to MI remains uncertain.

**ogm[GRCh38] der(13)(pter→q31.1::q31.1→q31.1::q31.1→q31.1::q31.1→ter)(pter_80,678,653~80,715,410::83,913,649_83,509,040::80,179,852_80,678,653::80,678,653~80,715,410_qter)**

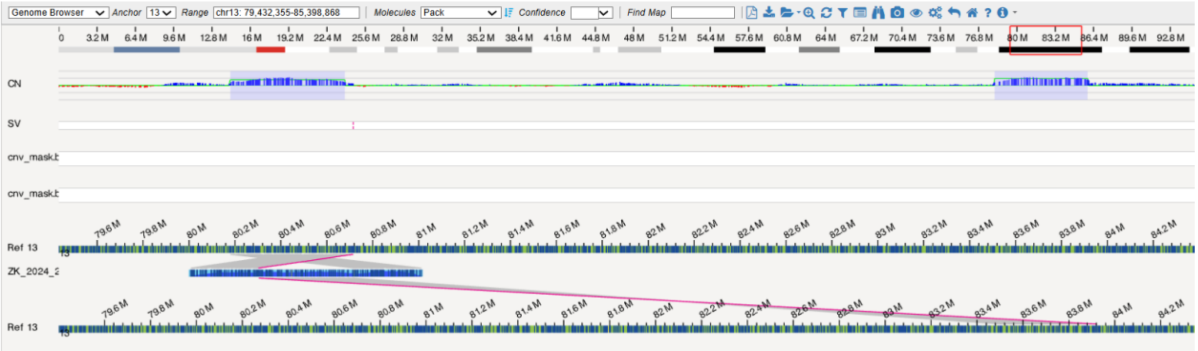


The complex variant, ogm[GRCh38] der(13)(pter→q31.1::q31.1→q31.1::q31.1→q31.1::q31.1→ter)(pter_80,678,653~80,715,410::83,913,649_83,509,040::80,179,852_80,678,653::80,678,653~80,715,410_qter), consists of a rearrangement of a 903.4 kb of genomic material in a region 3.7 Mb in size, involving an inversion and insertion of a region that contains the *SLITRK1* gene (OMIM:609678) (three copies total), and a duplication containing the *SPRY2* gene (OMIM:602466) (three copies total), that may originate from an insertional translocation or recombination event. To the best of our knowledge, none of the genes involved have been implicated in human disease in three copies. The insertion site of the inverted duplication did not contain genes. No similar rearrangements were found in any of the control databases. The significance of this finding remains unknown.

**ogm[GRCh38] 15q21.3(55,978,853_56,068,020)×1**

A

B

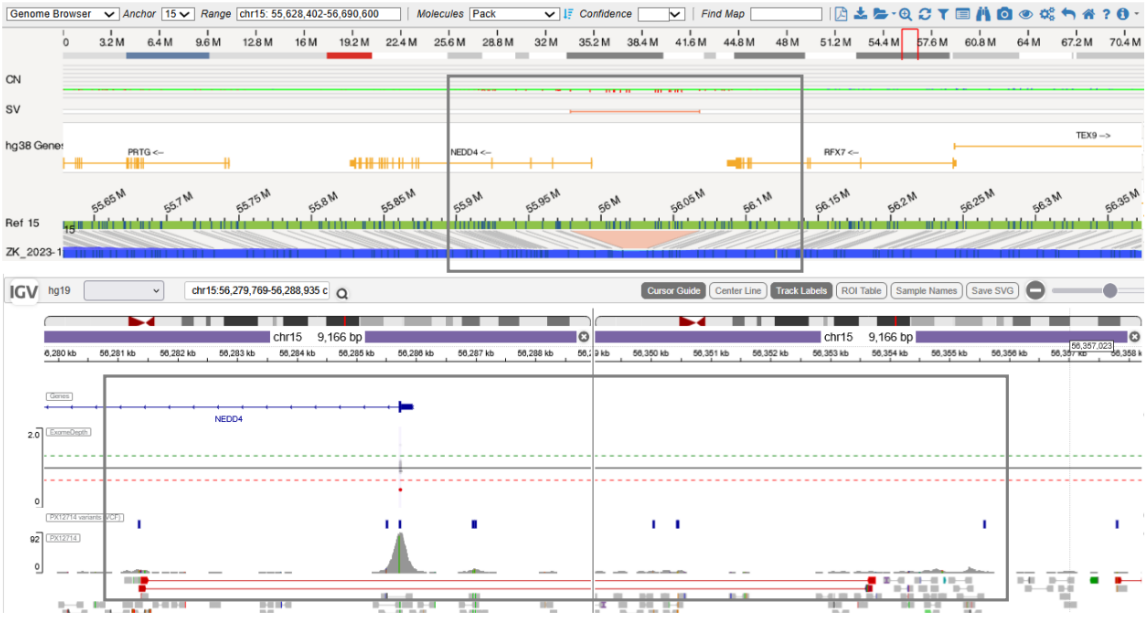


The ogm[GRCh38] 15q21.3(55,978,853_56,068,020)×1 is a 72.1 kb heterozygous deletion in the 89.2 kb region, that includes first exon of the *NEDD4* gene (NM_006154.4). *NEDD4* (OMIM:602278) is involved in cell differentiation during spermatogenesis (*14*), and its expression is lower in patients with asthenozoospermia (*15*).

**ogm[GRCh38] 17p13.1(9,624,197_9,638,372)×1**

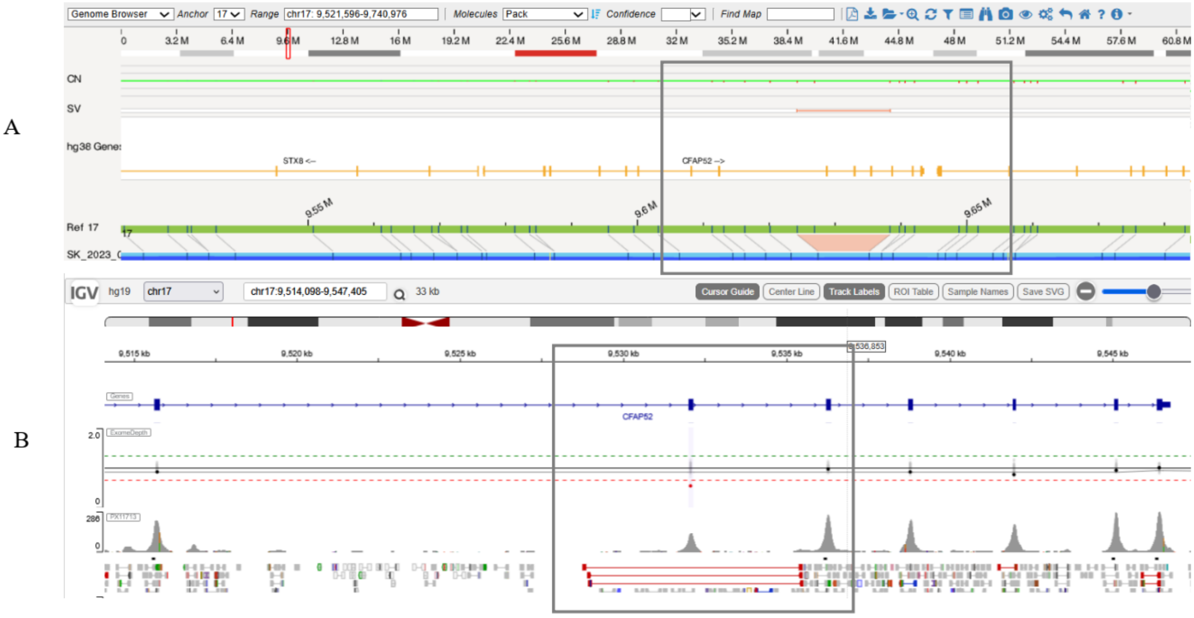


The 6.4 kb deletion ogm[GRCh38] 17p13.1(96,24,197_9,638,372)×1 in located in the 14.2 kb region, containing exons 9, 10 and 11 of the *CFAP52* gene (OMIM:609804, NM_145054.5). Biallelic pathogenic variants in the *CFAP52* gene (OMIM:609804) represent a known cause of heterotaxy with MI (OMIM:619607, (*16*)), and deletions of other exons have been described as causative (*17*). WES confirmation showed the deletion includes exon 9 of *CFAP52* (Supplement). Exon 9 is asymmetric, leading to a shift in the reading frame and thus a loss of the function of the protein that the gene encodes, however no additional SNV variants were shown by WES. So far two similar deletions have been described in the GnomAD database in individuals with unknown fertility status.

**ogm[GRCh38] 17p13.2(4,737,120_4,747,846)×0**

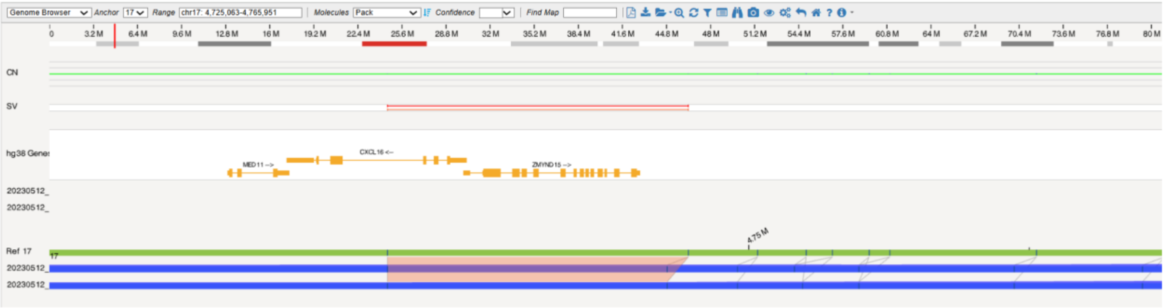


The homozygous deletion, ogm[GRCh38] 17p13.2(4,737,120_4,747,846)×0, was the smallest SV of interest detected, at 753 bp in size, and was present within 10.7 kb region containing the complete *ZMYND15* gene (OMIM:614312). Biallelic loss-of-function pathogenic variants in the *ZMYND15* gene have previously been associated with spermatogenesis disorder 14 (OMIM:615842)(*18*). The exact location of the deletion could not be determined due to the low DLE-1 resolution in this region and the SV could not be located using WES sequencing.

**ogm[GRCh38] 17p12(11,815,944_11,846,888)×1**

A

B

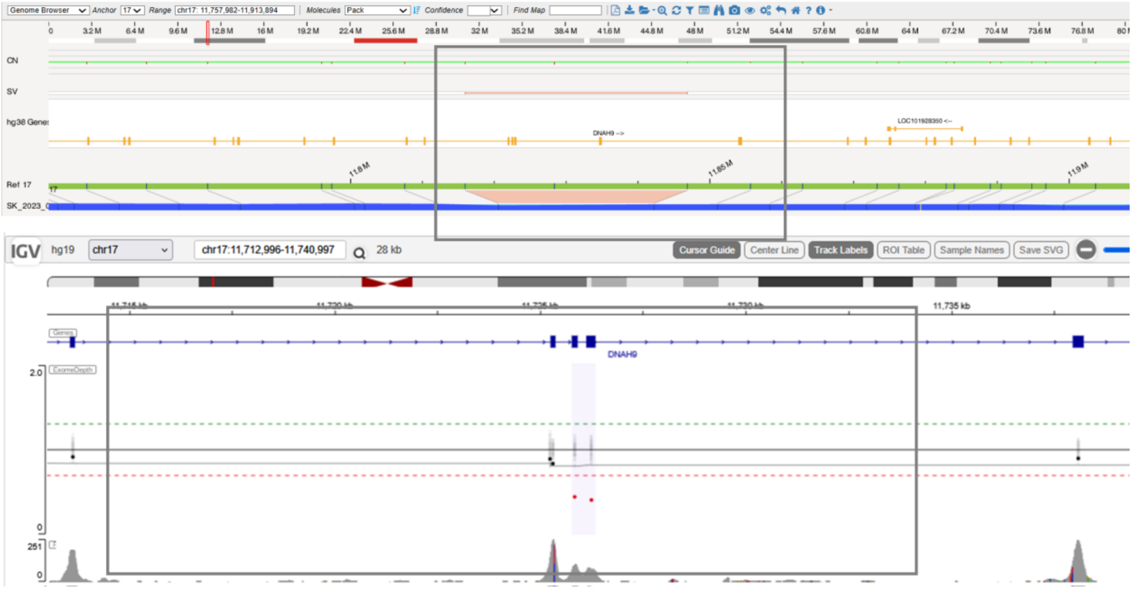


The 9.3 kb deletion ogm[GRCh38] 17p12(11,815,944_11,846,888)×1 is located in the 30.9 kb region containing exons 46 to 49 of the *DNAH9* gene (NM_001372.4). Biallelic pathogenic variants in the *DNAH9* (OMIM:603330) gene represent a known cause of primary ciliary dyskinesia type 40, which is associated with azoospermia and MI (OMIM:618300)(*19*). While WES confirmation showed the exons 47 and 48 are involved in the deletion, however the second SNV variant was not found.

**ogm[GRCh38] 18q21.1(46,124,191_46,130,285)×1**

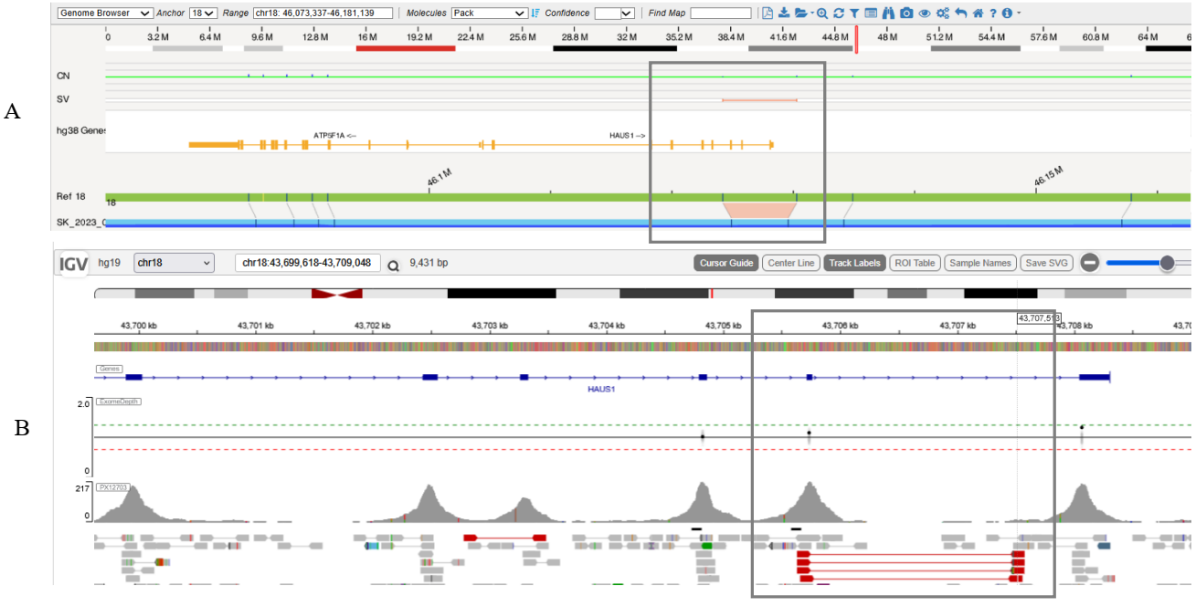


The 1.4 kb deletion ogm[GRCh38] 18q21.1(46,124,191_46,130,285)×1 was located in the 6.1 kb region containing exons 7-9 of the *HAUS1* gene (NM_138443.4). *HAUS1* (OMIM:608775) is involved in the mitotic spindle assembly and maintenance of chromosome integrity and is a candidate MI gene (*20*). This variant was one of the rare small SV also found in the comparison cohort. The deletion is not present in the internal Bionano database, while some overlapping deletions are present in the DGV and GnomAD databases, which contain data from healthy subjects with unknown fertility status. WES confirmed this variant to involve the exon 7 of the *HAUS1* gene.

**ogm[GRCh38] inv(20)(q11.23q13.12)(36,941,845_45,602,034)**

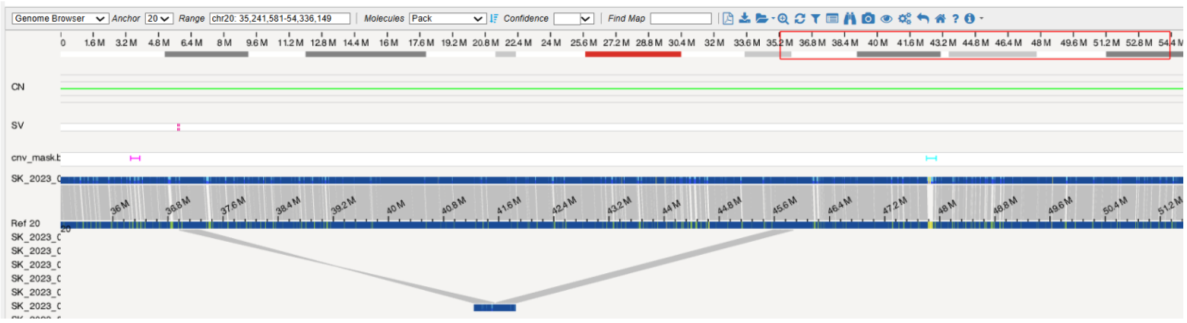


The largest autosomal SV we identified was a heterozygous paracentric 8.6 Mb inversion on chr20 (ogm[GRCh38] inv(20)(q11.23q13.12)(36,941,845_45,602,034)) that included over a hundred genes. While balanced inversions generally do not affect the health of the carrier, such changes may lead to unbalanced changes in gametes due to an inappropriate process of homologous recombination (*21*). To the best of our knowledge, chromosome 20 inversions have not been described in infertile men, however larger pericentric balanced inversions and rare paracentric balanced inversions of other chromosomes have been previously described in MI (*22*–*25*). The absence of paracentric inversions of the smaller chromosomes from the literature may reflect the difficulty in visualizing the banding pattern.

**ogm[GRCh38] Xq21.2(85,357,190_85,364,357)×0**

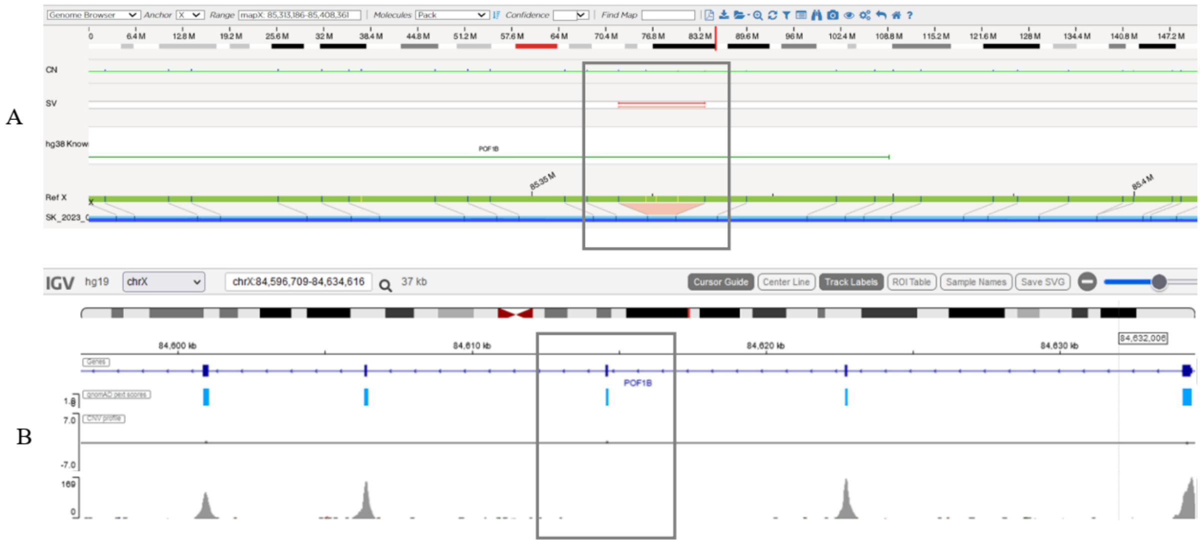


The homozygous deletion ogm[GRCh38] Xq21.2(85,357,190_85,364,357)×0, involves the symmetric exon 4 of the *POF1B* gene (OMIM:300603, NM_024921.4)(A). The deletion was confirmed using WES (B). While biallelic pathogenic variants in the *POF1B* cause premature ovarian failure in women (OMIM:300604), the role of this testis-expressed gene in men remains unknown.

**ogm[GRCh38] Xp22.31(8,796,917_8,802,256)×0**

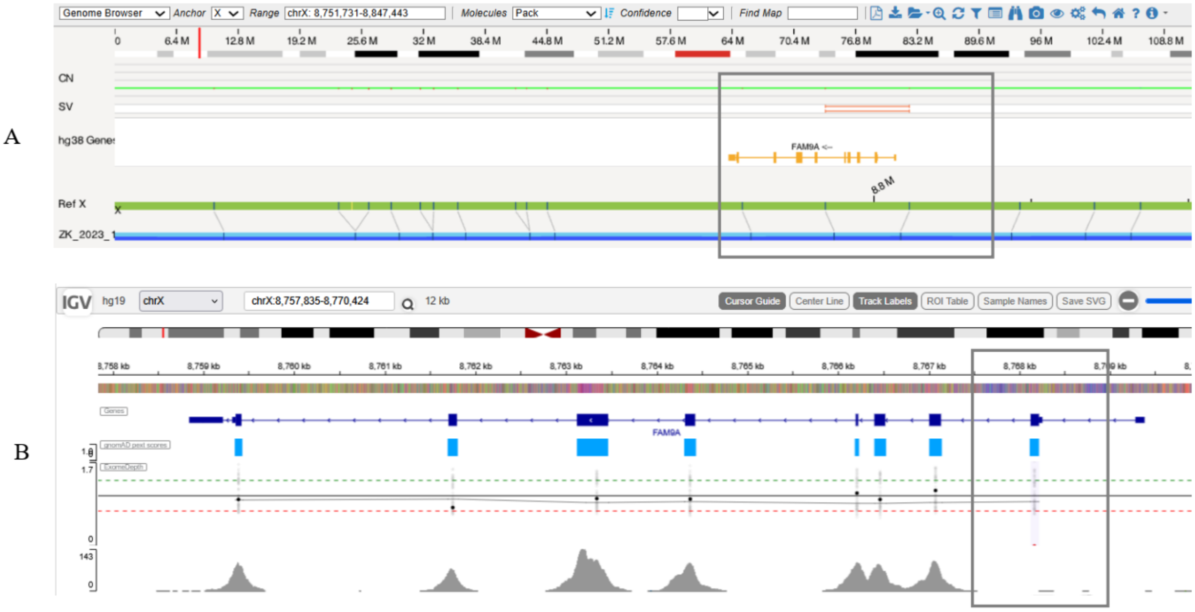


The ogm[GRCh38] Xp22.31(8,796,917_8,802,256)×0, involves exons 1-5 of the *FAM9A* gene (OMIM:300477, NM_174951.3)(A), and was confirmed as located in the exon 2 of the *FAM9A* gene by WES (B). *FAM9A* is exclusively expressed in the testes (*26*, *27*), but its role in MI has not been finally resolved (*28*).

**ogm[GRCh38] inv(X)(q28q28)(149,561,862_149,917,629)**

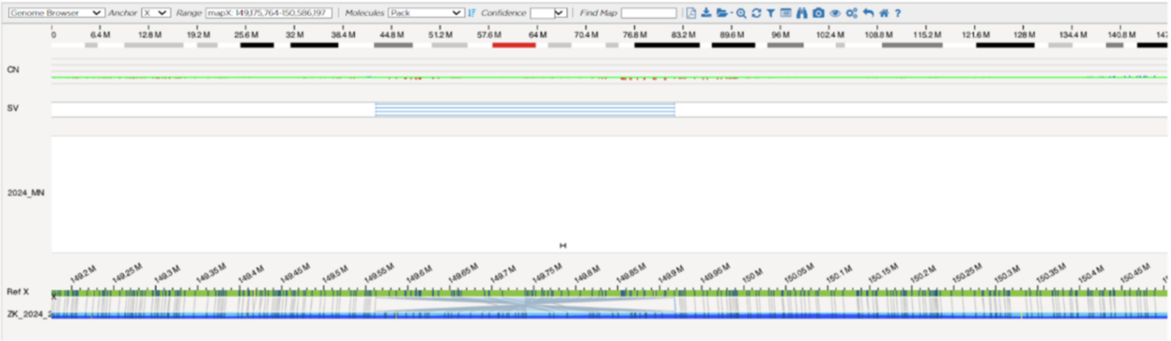


We identified a 355.7 kb hemizygous inversion ogm[GRCh38] inv(X)(q28q28)(149,561,862_149,917,629) in two patients (Table 1). This inversion included genes *MAGEA9B* (OMIM:300764), *FAM11A* (OMIM:300031), *MAGEA11* (OMIM:300344), *MAGEA9* (OMIM:300342), *LINC0085* (OMIM:300892), and *MAGEA8* (OMIM:300341), that belong to the family of testicular antigens (*29*). Based on animal studies, they are said to influence the correct development of the testes (*30*), but their role in MI is not definitively explained. The resolution of OGM did not allow us to confirm the presence of a previously reported intergenic MI-associated deletion (CNV67) in the inversion region (*31*).

**ogm[GRCh38] Xq24(120,866,580_120,990,165)×10, Xq26.3(135,238,491_135,743,639)×3**

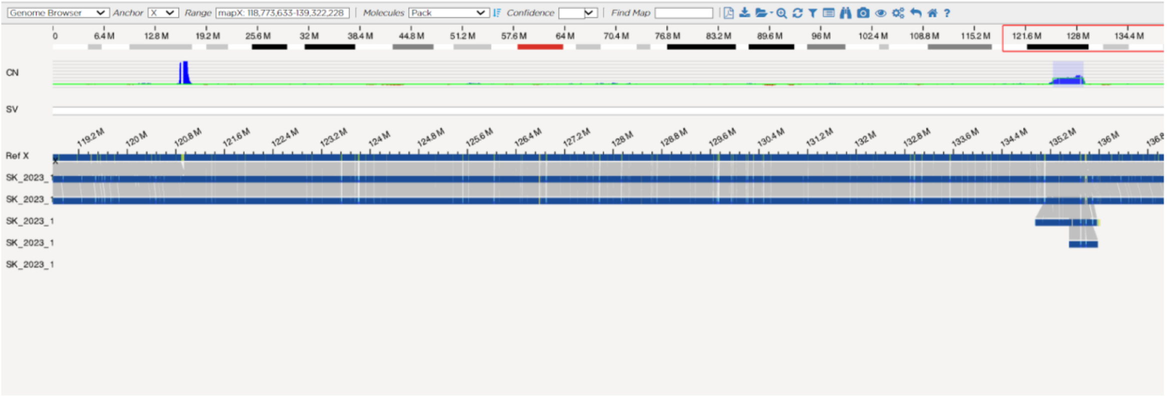


Two presumably hemizygous amplifications of interest on chrX were detected in a single patient, ogm[GRCh38] Xq24(120,866,580_120,990,165)×10, and Xq26.3(135,238,491_135,743,639)×3. Previously, chrX microduplications, have been shown to be enriched in MI cohorts (*32*, *33*). While the Xq24 region overlapping our SV is frequently present in two copies in the general population (ClinGen ISCA-46733), our patient had approx. 10 copies. The detected amplification in Xq26.3 contains *ZNF449* (OMIM:300627) and *ZNF57D* (OMIM:314997), the function of which is unknown, but none of the overlapping CNV in these genes found in the Decipher database were paternally inherited. Smaller overlapping duplications have been described previously in MI (*12*, *32*). In the GnomAD database, we found 7 male individuals with unknown fertility status, with an overlapping complex duplication (DUP_CHRX_EB12C2D5) that additionally includes the *CT55* gene (OMIM:301105), which was previously associated with a defect in spermatogenesis. In our patient, the *CT55* was not included in the amplification region, however, the indirect influence of amplification on its expression cannot be excluded.

ChrY microdeletions (Table 1)

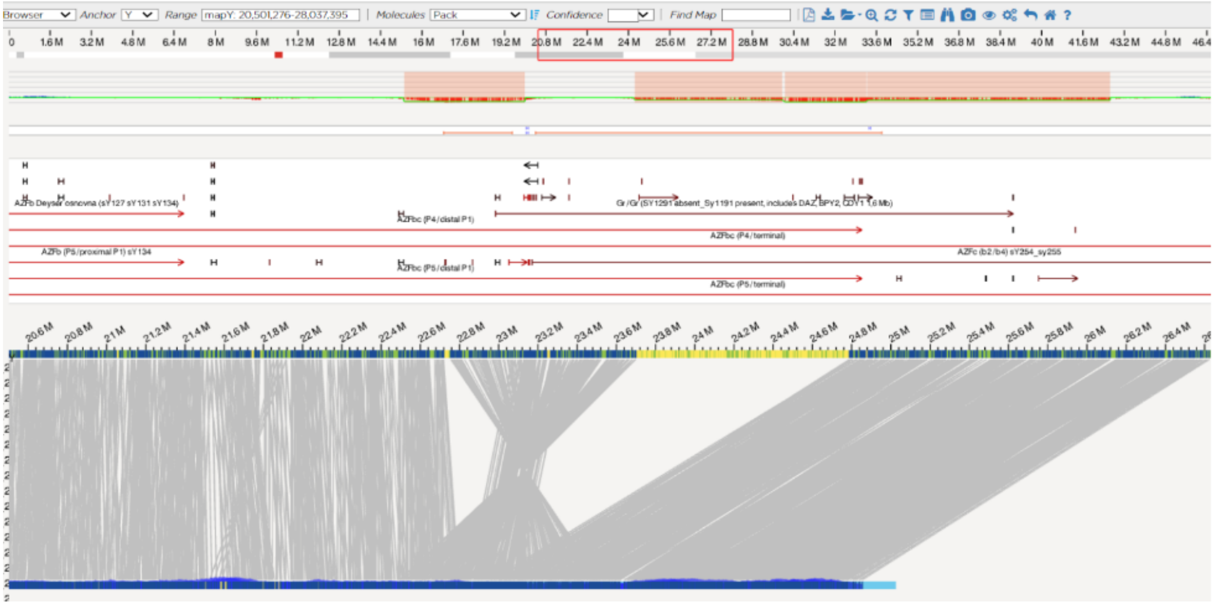

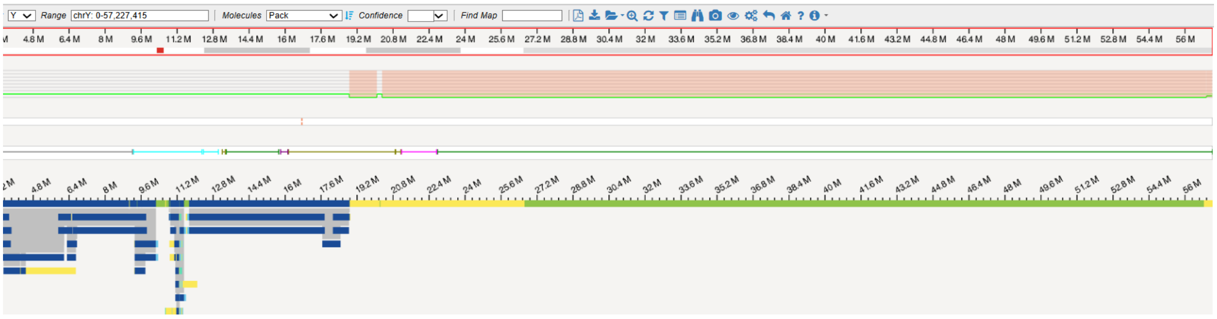

Different SV on chrY were the most common finding in our cohort. While the chrY microdeletion are a known, routinely tested cause of MI (Fan and Silber, 2002; Navarro-Costa *et al.*, 2010; Yu *et al.*, 2015; Colaco and Modi, 2018), OGM revealed their variability and complexity is much greater than what can be determined using PCR approaches, as further discussed below. Initially, the SV identified on chrY using OGM were missed by the classic chrY microdeletion testing approach that relies on molecular probes (for details on probes please see Methods).

This is probably because the high DNA repetitiveness on chrY masks the underlying complexity of SV (e.g. basic molecular probe binding sites may occur in several copies, and only full deletions of all copies are apparent as deletions). Therefore, classic chrY microdeletion testing is unlikely to do justice to the true structural complexity associated with chrY.

OGM showed several chrY SV to be more complex than indicated by the classical testing approaches. Among them are the AZFbc deletions, which are relatively big, but are known to not necessarily lead to complete azoospermia (Repping *et al.*, 2002; Krausz *et al.*, 2014), and have previously been described in the context of hypospermatogenesis (Costa *et al.*, 2008). Furthermore, the AZFc type b2/b3 deletions were previously described in individuals with reduced fertility (Ferlin *et al.*, 2005; Wu *et al.*, 2007; Eloualid *et al.*, 2012; Bansal *et al.*, 2016; Zhou *et al.*, 2019), but their definitive role in MI remains unclear in the European population (Repping *et al.*, 2004; Eloualid *et al.*, 2012). It is unclear if and what is the biological basis of such differences between different populations, however these differences may be inversion or haplotype based and with better resolution of chrY in the future, we may begin to understand the underlying cause of such differences.

By subsequently using the extended version of the chrY microdeletion assay (Devyser AZF Extension kit), we were able to retrospectively confirm almost all the SV identified using OGM, such as the AZFb (azoospermia factor b) deletions with P4-type breakpoints (Repping *et al.*, 2002; Costa *et al.*, 2008; Krausz *et al.*, 2014), AZFc (azoospermia factor c), and smaller AZFc type b2/b3 breakpoints (absence of sY1192, and sY1191) (Rozen *et al.*, 2012), and AZFc gr/gr breakpoints (absence of sY1291, presence of sY1191) (Repping *et al.*, 2003; Masoudi *et al.*, 2016) (Table 1) (please see Methods section and Supplement for further details and mapping of the Devyser probes).
