## Supplementary material for "The landscape of structural variants in male infertility identified by optical genome mapping": SI Table 1

Table S1: SV identified in MI patients and controls.

| Chromosome region | Nucleotide coordinates GRCh38 | Size, kb | Abnormality type | Reported genes / region | Copy number | zygosity | Patients (n = 88) | Controls (n = 132) |
|---|---|---|---|---|---|---|---|---|
| **Yq11.223** | 23,008,278_23,494,114 | 485.8 | deletion | AZFc gr/gr risk factor | 0 | hemi | 1 | 1# FT |
| **Yq11.223** | 23,071,190_23,731,504 | 660.3 | deletion | AZFc gr/gr risk factor | 0 | hemi | 1 | 1# RM |
| Yq11.223q11.23 | 22,563,555_26,033,310 | ~3,469.8 | duplication | AZFc b2/b4 | 2 | hemi | 2# | 5# RM |
| Yq11.223<br>Yq11.23 | 22,893,762_23,413,097<br>25,628,806_26,273,822 | 519.3<br>645.0 | duplication | AZFc b2/b4 partial | 2 | hemi | 1 | 1#FT, 1#RM |
| 10q11.22 | 46,100,908_46,680,405 | ~579.5 | amplification | *PPRY1* (OMIM:601790), *GPRIN2* (OMIM:611240), *SYT15* (OMIM:608081), *ANTXRL* (HGNC:27277) | >4 | / | 5 (1*) | 1# RM |
| 18q21.1 | 46,124,191_46,130,285 | 1.4 | deletion | *HAUS1* (OMIM:608775, NM_138443.4) | 1 | het | 1 | 2 |
| 6p21.32 | 32,354,608_32,357,248 | 2.3 | insertion | *TSBP1* (OMIM:618151, NM_001286474.2) | / | hom | 1 | 0 |
| | | | | | / | het | 6 | 9 |
| 12q23.1 | 99,598,042_99,615,914 | 15.2 | deletion | *ANKS1B* (OMIM:607815, NM_001352186.2) intronic | 0 | hom | 1 | 0 |
| | | | | | 1 | het | 1 | 1 |
| 17p13.2 | 4,737,120_4,747,846 | 0.753 | deletion | *CXCL16* (OMIM:605398) in *ZMYND15* (OMIM:614312) | 0 | hom | 1 | 0 |
| | | | | | 1 | het | 1 | 1 |
| | | | | | | **total** | 22 | 23 |

*Patients with more than one SV of interest, # overlapping variant, FT-father from rare-disease trio cohort, RM-healthy male from recurrent miscarriages cohort. Hemi – hemizygous, het – heterozygous, hom – homozygous.
